# Empirical non-linear modeling & forecast of global daily deaths of COVID-19 pandemic & evidence that a “third wave” is beginning to decay

**DOI:** 10.1101/2020.07.07.20147900

**Authors:** Jorge Sánchez-Sesma

## Abstract

**Objectives:** The present COVID-19 pandemic (C19P) is challenging our socities all over the world. In this work, based on massive health information daily updated, the C19P daily death numbers at a global level, are modelled, analyzed and forecasted.

**Methods:** Two empirical models are proposed to explain daily death (DD) records: a) self-similar (SS) recurrences of the global responses, and b) geometric averaging of two independent SS models for global DD records.

**Findings:** The detected self-similar recurrences in the global response suggest three global “self-similar waves” that support multi-month forecasts of the DD numbers. However, there are upper and lower-limit SS forecast DD scenarios that were jointly integrated with a geometrical average (GA) model, that support the existence of a moderated “third wave”, with a decaying stage for the next months (July-September 2020). It appears that the “third world” (South America [SAM]+Asia [ASI] +Africa [AFR]), is the actual “big player”, (following China, and Europe [EUR]+North America [NAM]) with its biggest contribution to a global “third wave” (W3) of C19P.

**Conclusion:** The empirical global modeling of the C19P has suggested us a possible moderated W3 scenario, with contributions mainly coming from the third world people. This moderated W3 scenario, after to be calibrated with the last weeks, has provided to stakeholders of significant data and criteria to define, sustain and support plans for the next months, based on data and self-similarities. These scenarios provide a well-based perspective on non-linear dynamics of C19P, that will complement the standard health and economic models.

## INTRODUCTION

> *“With no vaccine to protect against influenza infection and no antibiotics to treat secondary bacterial infections that can be associated with influenza infections, control efforts worldwide were limited to non-pharmaceutical interventions such as isolation, quarantine, good personal hygiene, use of disinfectants, and limitations of public gatherings, which were applied unevenly*.*”*
>
> ***1918 Pandemic (H1N1 virus)*** *(https://www.cdc.gov/flu/pandemic-resources)* [1]
>
> *“In December, 2019, Wuhan, Hubei province, China, became the centre of an outbreak of pneumonia of unknown cause, which raised intense attention not only within China but internationally. Chinese health authorities did an immediate investigation to characterize and control the disease, including isolation of people suspected to have the disease, close monitoring of contacts, epidemiological and clinical data collection from patients, and development of diagnostic and treatment procedures*.*”*
>
> ***A novel coronavirus outbreak of global health concern***
>
> *Wang et al. (2020)* [2]
>
> *“The extremely rapid spread of the virus across the globe began in the most economically developed regions where international trade and business is prioritized. After initially following the corridors and international trade routes between developed countries, the virus spread later to developing countries*.*”*
>
> ***Economic globalization and the COVID-19 pandemic: global spread and inequalities***
>
> *(Jeanne et al*., *2020)*[3]

In 2020, the COVID-19 pandemic has unprecedently affected health’s people and enhancing inequalities (Jeanne *et al*., 2020) [3], but also has contributed to global recession with adverse consequences for unemployment and poverty (IMF/WB, 2020)[4].

This extreme situation, demands stronger and efficient states that provide a “healthy” balance of the give-and-take relationship between the state and the individual, offering the benefits of the accumulated advances of science, technology and social sciences to the lay people (*The Lancet*, 2020) [5].

It is undeniable that we are in the midst of health, environmental and economic global crisis. The adequate modeling of these widespread phenomena which belongs in the Earth system dynamics requires precise knowledge of two kinds of internal processes: those operating in the physical, chemical and biological systems of the planet and those occurring within its human societies, their cultures and economies (Donges *et al*., 2020) [6].

A common characteristic of the complexities of all these processes is self-similarity, or the property of having a substructure analogous or identical to an overall structure in their dynamics. However, since 1987, self-similarity was identified as a part of self-organized criticality systems (Bak et al. 1987, 1988)[7,8]. These systems, that range from the physical to the biological to the social, have provided new concepts, particularly in the field of physiology and medicine. For instance, it is now recognized that fractal architecture of patterns serves as robust, stable structures for the regulation and maintenance of vital functions of lungs, heart, liver, kidneys, brain, etc. (Selvam, 2017) [9].

In this work, simple self-similar models are applied to the past and forecasted evolution of the pandemic of C19P, and their results were non-linearly averaged providing an explanation for the past month and suggesting future decaying trends for the next months.

Our societies and their stakeholders require continuously updated and verified global scenarios. Here we have provided, self-similar scenarios, with an objective and well-based perspective on data and dynamics of C19P, that will complement the standard health and economic models. In what follows, the data, models, results, discussion and preliminary conclusions are presented.

## METHODS

### Data

The number of daily deaths (DD) for the world and six continents (North America, South America, Europe, Africa, Asia and Oceania) [WLD, NAM, SAM, EUR, AFR, ASI & OCE; hereafter], due to C19P were obtained from Oxford-OWID, (2020) [10] (Actualization date: July 5th, 2020). They are displayed in Figure 1, and graphs of DD for two countries, China and Belgium, are also displayed in the same Figure. The abcissa indicates time in the continuous numbering of days, the Julian day (JD), of 2020.

**Figure 1.**
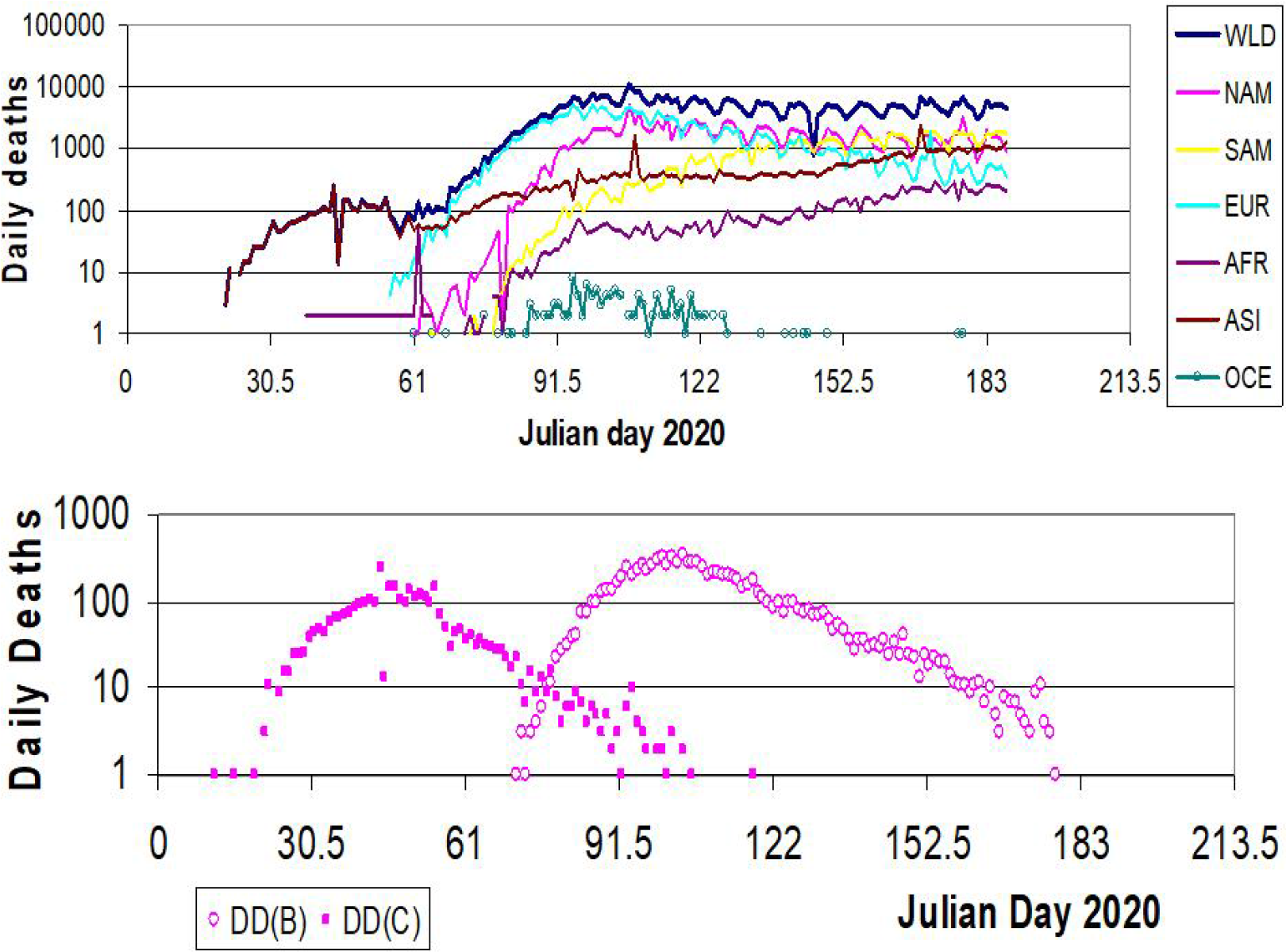
Comparison of the C19P DD records. a) DD records for the world and six continents (North America, South America, Europe, Africa, Asia and Oceania) [WLD, NAM, SAM, EUR, AFR, ASI & OCE, respectively); b) China and Belgium, DD(C) and DD(B), records. Data is from Oxford-OWID. Julian Days 61, and 122 represent March 1^st^, and May 1^st^, respectively.

### Models

A self-similar model, *modDD*(*t*), is defined as:

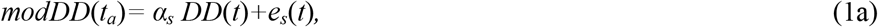

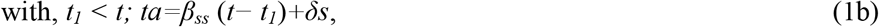

where: *t*_*a*_ is the adjusted time for the self-similar process, *α*_*s*_ is the value amplification factor, *β*_*s*_ is the time amplification factor, *δ*_*s*_ is the lag of the self-similar process, and *t*_*1*_ is the initial time for the analysis and the modeled periods, respectively.

This self-similar model is evaluated with a minimization of errors. The parameters *t*_*1*_, the lag *δ*, the amplification factors of values and time scale (*α* and *β*) are estimated through an iterative process that minimizes the standard deviation of the self-similar error, *e*_*s*_(*t*).

A geometric average model, *gmodDD*(*t*), that integrate the values of two SS models (*mod1DD*(*t*) and *mod2DD*(*t*)), is defined as:

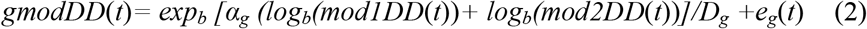

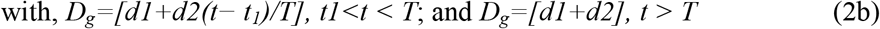

where: *t*_*1*_ is the initial time for the geometrical process, *α*_*g*_ is the value amplification factor (around 0.5), *D*_*g*_ is the linear decaying factor, d1 is its initial value, and d1 is its complementary final value and *T* is its time scale.

This nonlinear model is evaluated with a minimization of errors. The values of *t*_*1*_, *T* and the amplification factor *α*_*g*_ are estimated through an iterative process that minimizes the standard deviation of the error, *e*_*g*_(*t*). The Standard deviation, provides information to estimate a corresponding ranges of values of the *gmodDD* forecasts.

## RESULTS

Based on data gathered by Oxford-OWID (2020) [10], the concentration of deaths in North America (NAM), South America (SAM) and Europe (EUR) is analyzed. With only 23 % of the global population these three continents NAM+SAM+EUR, together, have concentrated more than 87% of the global accumulated deaths.

However, the comparison is more extreme when the multi-continental average of deaths per million are taken into account, with values of 241 and 7.5, for the macro-continents NAM+SAM+EUR and AFR+ASI+OCE, respectively. The corresponding deaths ratios indicate that the NAM+SAM+EUR values are 32.2 times higher than those for AFR+ASI+OCE. Additionally, in order to provide a comparison of daily changes, estimations based on average multi-continental values over the last two weeks are the following: DD changes are −2.62 and 0.15 %, showing decrease and grow, for the macro-continents NAM+SAM+EUR and AFR+ASI+OCE, respectively.

After developing this spatial distribution analysis of C19P, two self-similar models, proposed herein (See Eq. 1), were adjusted to the global daily death records DD(G) of C19P. The results are shown in Figure 2.

**Figure 2.**
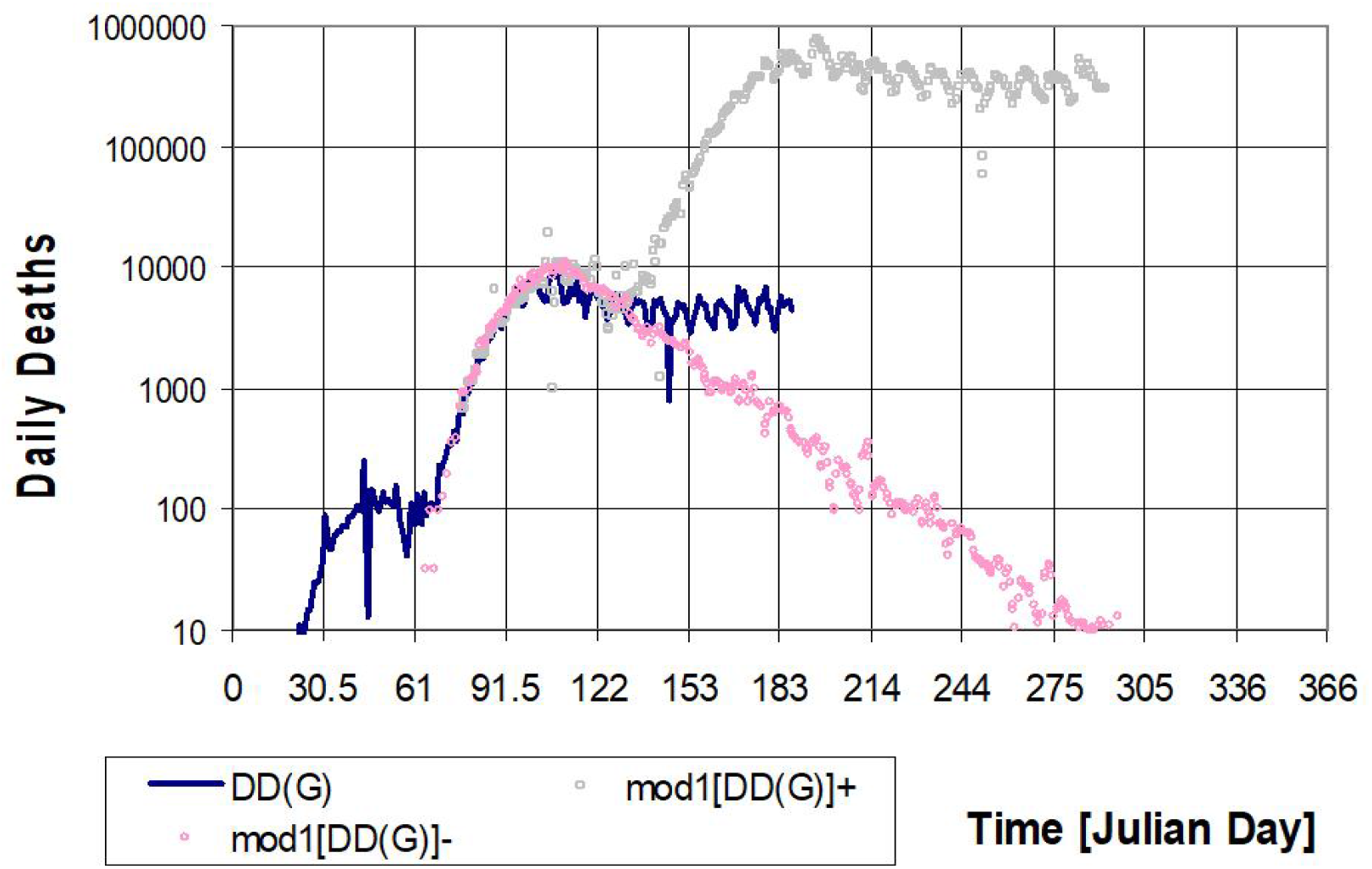
Global daily death [DD(G)] records. The DD(G) record and its self-similar modeling for the Julian days 61 to 290 of 2020. Two models mod1[DD(G)]+ and mod2[DD(G)]-, upper- and lower-limit models, based on the DD(G) [βs =1.42, δ s = 42 days, αs=75] and DD(B) [βs =1.42, δ s = −38 days, αs =31] records, respectively. Note that Julian days 91 and 122 represent April 1^st^, and May 1^st^, respectively

One model, mod1[DD(G)]+, that is based on the DD(G) record (*βs* =1.42, *δs* = 42 days, *αs*=75), explains more than 52% of the variance of the same record from JD 75 to 132, however after JD 132, it represents a global upper-limit scenario for the W3, or a “business as usual” (BAU) scenario, because it implies the limited and “soft” social-rules applied to content the C19P. The other model, mod2[DD(G)]-, that is based on the DD record of Belgium (DD(B)) (*βs*=1.42, *δ s*=-38 days, *αs*=31), also explains with more than 76% of the variance the DD(G), from JDs 75 to 132. However after JD 132, in its decaying phase, it represents a global lower-limit scenario for the W3, the restricted conditions applied (although late) in Belgium (and Western Europe).

Figure 3 shows the two self-similar models, Mod[DD(G)]+ and Mod[DD(G)]-, and its corresponding scenarios, but also it shows a geometric average of those models, geomDD(G), based on Eq. 2. After its calibration and evaluation of the standard deviation of errors, this geometrical model (*αg*=0.45, *d1*=0.6, *d2*=2.4, *T*=50 days), that explains with more than 60% of the variance the DD(G), from JDs 146 to 184, represents a decaying scenario for the next months.

**Figure 3.**
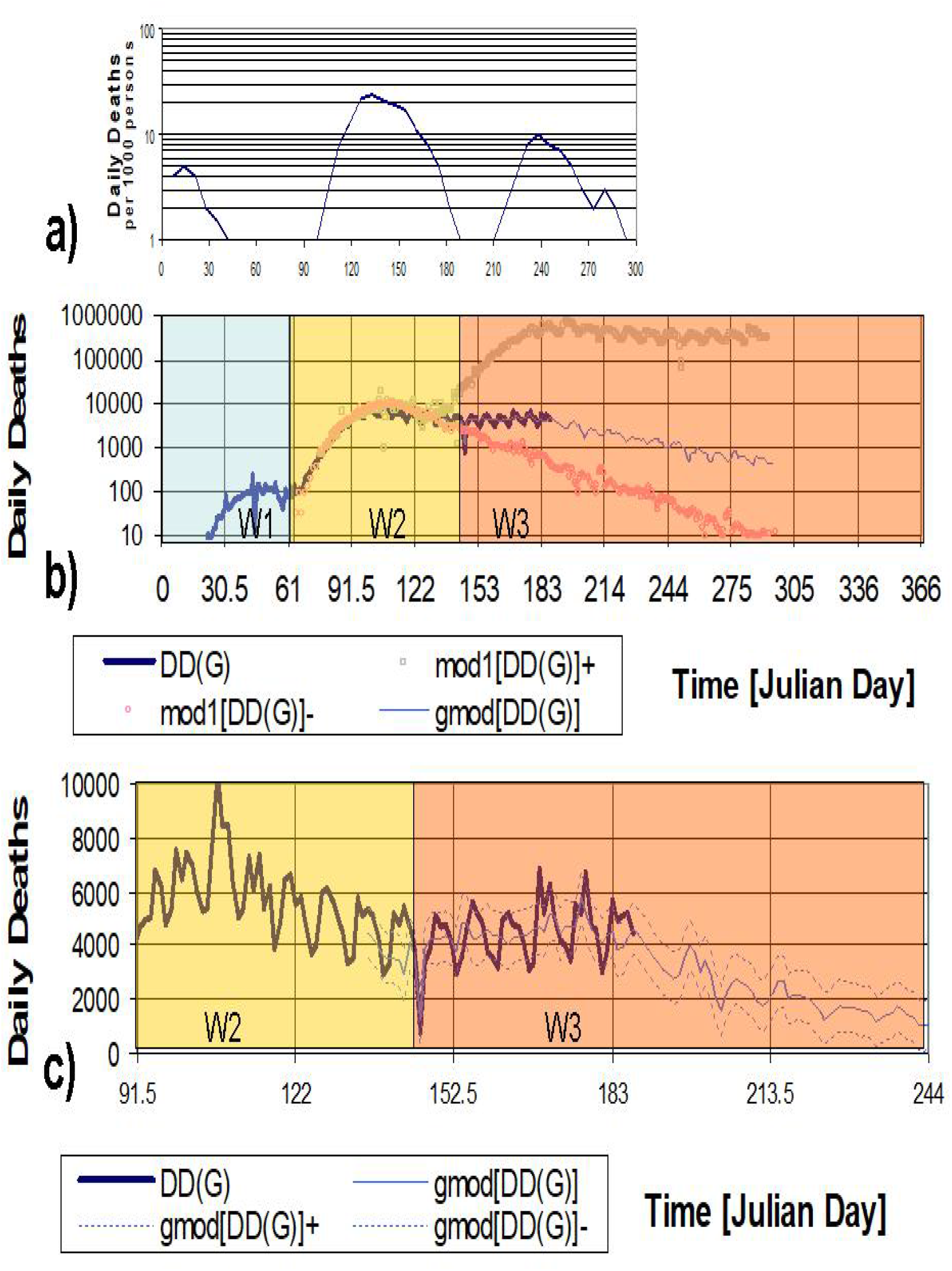
Comparison of London daily death records of 1918 influenza and global death CoViD-19 records and models. **a)** London records of deaths per 1000 persons (abcissa 0 indicates June 6^th^ 1918), **b)** SS modelling and forecast of DD(G) (Figure 2), and their geometric average gmod[DD(G)], adjusted to the DD(G) record over the last month (αg=0.45, d1=0.7, d2=2.0, T=50days) and its forecasts for the next three months; and **c)** a zoom of b) indicating a range (+/-StdDev) of the gmod[DD(G)]. Three waves of the C19P are also displayed (W1, W2, W3). Julian days 91 and 122 represent April 1^st^, and May 1^st^, respectively.

In the same Figure 3, it has been enhancing the occurrence of three “waves” (W1,W2 and W3). The “Wave 1” (W1) is associated with the Chinese outbreak early in 2020. The “Wave 2” (W2) is associated with the European and North American outbreaks early in 2020 but is replicated by a lagged and non-linearly adjusted Belgian record. The “Wave 3” (W3), after the JD 165, the DD(G) represents a forecasted upper limit scenario of the third wave (W3), and the continuation of the self-similarly adjusted DD(B) record, or Mod[DD(G)]-, would be considered a lower limit scenario, if there were no other new countries to be considered important “players” of the pandemic.

In order to provide more elements about the potential existence of the W3, the records of continental daily death records of DD, for two multi-continents, has shown different increasing and decreasing trends. Figure 4, displays, the NAM+EUR-mex and SAM+AFR+ASI+OCE+mex records that emphasize different response dynamics to the C19P. The increasing trend of the “third world” represented by the integrated contributions of SAM+AFR+ASI+OCE+mex, appears to have gained its maximum and surpassed the integrated contributions of NAM+EUR-mex.

**Figure 4.**
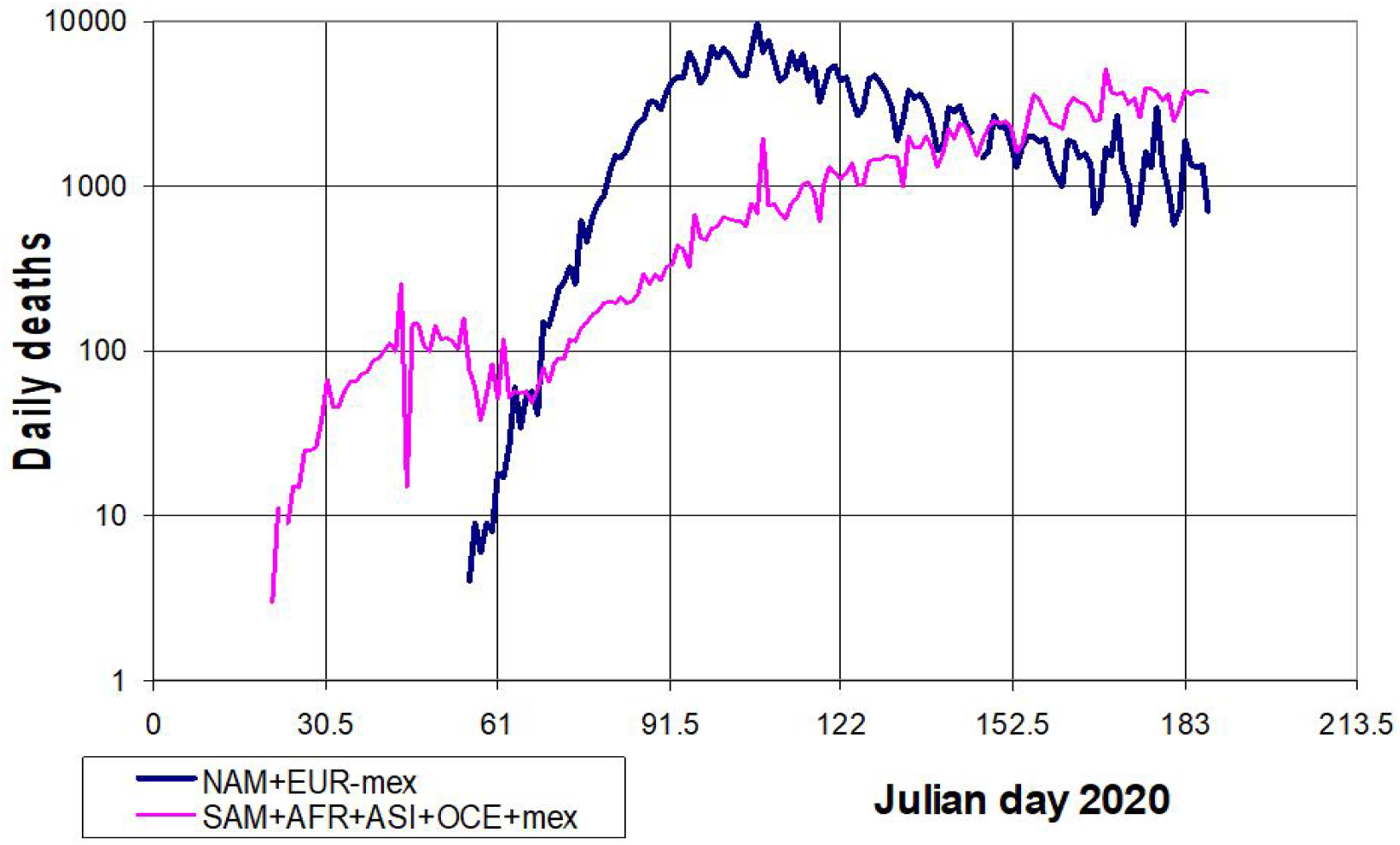
Daily death [DD] records for two groups of continents, the NAM+EUR-mex and SAM+AFR+ASI+OCE-mex. The adjusted models provide elements to follow the geo-center of the C19P. Julian Days 91 and 122 represent April 1^st^, and May 1^st^, respectively.

## DISCUSSION

Firstly in this discussion, it is important to emphasize the historical evidence of three “waves” of the 1918-1919 influenza epidemic that cause numerous deaths in London, UK. Figure 3 compares these three waves of that pandemic in London (Taubenberger and Morens, 2006)) [11], with the detected and forecasted waves for the C19P. It is important to mention that the separation of the C19P waves is around 60 days, much lower than the separation, of around 120 days, of the 1918-1919 pandemic waves registered at London. This difference indicates that the actual activities are faster than those of 1918-1919.

Empirical self-similar modeling of the C19P has shown us upper- and lower-limit forecasts related to “business as usual” (BAU) and strict measures like those taken in Belgium (and China), respectively. In this work, another empirical non-linear model (a geometric average) has shown us global and decaying trends that confirm the existence of a “third wave” but a more flattened one. As in London, early 1919, the world appears to be, as right now, in the middle of a W3.

Two facts support the continuation of a C19P “third wave”. Firstly, the irregular spatial distribution and dynamics of DD over the continents. Secondly, the forecast modelling of DD, that shows, several months in advance, a trend estimation of DD values that indicate a global decaying continuation (based increasingly on the “third world” contributions) for the next months.

The Director of the PAHO confirmed that in his latest message about C19P, from the May 26^th^ 2020 [12] and pointed out: “As global cases topped 5 million this past week, Latin America surpassed Europe and the United States in the daily number of reported coronavirus infections, numbers we suspect are even higher than we know. Two of the three countries with the highest number of reported cases are now in the Americas.”

## CONCLUSIONS

In 1918, within a global war, the WWI, the influenza spread all over the world and infected around 500 million people and cause deaths of around 50 million (CDC, 2020) [1]. In 2020, one century later, the pandemic threat, is back again with regional wars, a world economic war and a severe environmental crisis.

In this work, the 1918 influenza pandemic was seriously considered, because it was the most severe in recent history [1]. It should be noted that London, as the economic and cultural center of the world at that time, was affected by three “waves” of influenza deaths, that were registered during 1918-1919.

Taking into account the 1918 pandemic experience and the actual and amazing flow of medical information of the C19P from all over the world, it is proposed, analyzed and extrapolated self-similar models of C19P. The consequent results provide global death forecasts for the next three months that are based on non-linear and lagged modeling that show the movement of the center of activity from China to NAM and EUR (in March 2020), to other regions of SAM, ASI and AFR (in late May & June 2020). However, it is important to evaluate and discuss the C19P global evolution or “path” during the following weeks (July 6^th^ -July 18^th^), because this period will confirm us if the C19P is going to decline as modelled, constituing a favorable scenario for the global population, off course, with different regional impacts.

The actual health crisis of CoViD-19 pandemic is confronting the global society, however, this challenge in health, economy, and social practices, also offers opportunities to better change all these activities, through knowledge from accurate data and its analysis. Although CoViD-19 pandemic is severely affecting the health and economy of our societies all over the world, we are able to surpass this challenging situation with reasonable costs based upon the accumulated knowledge and resources channeled through our social institutions.

## Data Availability

Information used can be accessed in an open site

## ACKNOWLEDGMENTS

Thanks to the encouraging comments about the first versions of this work coming from Fabiola Mendez, Valeria Toledo, Jorge Aguirre, Sébastien Bourdin, Victor Toledo, Francisco José Sánchez-Sesma and Luis Pech.

## REFERENCES

1. CDC. Web page. (https://www.cdc.gov/flu/pandemic-resources): Retrived May 24th 2020.

2. Wang, C., Horby, P.W., Hayden F.G. and Gao GF, A novel coronavirus outbreak of global health concern, The Lancet, January 24th, 2020, doi:10.1016/S0140-6736(20)30185-9

3. Jeanne L, Bourdin S, Nadou F & Noiret G. Economic globalization and the COVID-19 pandemic: global spread and inequalities. [Preprint]. Bull World Health Organ. E-pub: April 23rd 2020. doi:10.2471/BLT.20.261099

4. COVID-19: The Regulatory and Supervisory Implications for the Banking Sector : A Joint IMF-World Bank Staff Position Note, IMF. Monetary and Capital Markets Department & World BankPublication, May 21st, 2020. https://www.imf.org/en/Publications/Miscellaneous-Publication-Other/Issues/2020-/05/20/COVID-19-The-Regulatory-and-Supervisory-Implications-for-the-Banking-Sector-49452

5. The Lancet, editorial May 2020.

6. Donges, J. F., Heitzig, J., Barfuss, W., Wiedermann, M., Kassel, J. A., Kittel, T., Kolb, J. J., Kolster, T., Müller-Hansen, F., Otto, I. M., Zimmerer, K. B., and Lucht, W.: Earth system modeling with endogenous and dynamic human societies: the copan:CORE open World–Earth modeling framework, Earth Syst. Dynam., 11, 395–413, https://doi.org/10.5194/esd-11-395-2020, 2020.

7. Bak, P., Tang, C., Wiesenfeld K.: Self-organized criticality: an explanation of 1/f noise. Phys. Rev. Lett. 59, 381–384. 1987.

8. Bak, P.C., Tang, C., Wiesenfeld, K.: Self-organized criticality. Phys. Rev. A. 38, 364–374. 1988

9. Selvam, A.M. Self-organized Criticality: A Signature of Quantum-like Chaos in Atmospheric Flows, Springer Atmospheric Sciences, ISBN 978-3-319-54546-2. 2017.

10. O-OWID. Oxford, Our World in Data. Retrived: July 5th 2020 https://covid.ourworldindata.org/data/ecdc/owid-covid-data.csv

11. Taubenberger JF, and Morens DM, 1918 Influenza: the Mother of All Pandemics, Emerg Infect Dis; 12(1): 15–22., doi: 10.3201/eid1201.050979, 2006

12. PAHO, (2020). Director’s opening remark. May 26th, 2020 https://www.paho.org/en/documents/weekly-press-briefing-covid-19-directors-opening-remarks-may-26-2020

